# Safety and effectiveness of RBD-specific polyclonal equine F(ab′)2 fragments for the treatment of hospitalized patients with severe Covid-19 disease: a retrospective cohort study

**DOI:** 10.1101/2022.04.07.22273558

**Authors:** Diego H. Farizano Salazar, Fernando Achinelli, Mariana Colonna, Lucía Perez, Analía A. Giménez, Maria Alejandra Ojeda, Susana N. Miranda Puente, Lía Sánchez Negrette, Florencia Cañete, Ornela I. Martelotte Ibarra, Santiago Sanguineti, Linus Spatz, Fernando A. Goldbaum, Carolina Massa, Marta Rivas, Mariana Pichel, Yanina Hiriart, Vanesa Zylberman, Sandra Gallego, Brenda Konigheim, Francisco Fernandez, Matías Deprati, Ian Roubicek, Diego H. Giunta, Esteban Nannini, Gustavo Lopardo, Waldo H. Belloso, EPIC Study Group

## Abstract

Passive immunotherapy has been evaluated as a therapeutic alternative for patients with COVID-19 disease. Equine polyclonal immunotherapy for COVID-19 (EPIC) showed adequate safety and potential efficacy in a clinical trial setting and obtained emergency use authorization in Argentina. We studied its utility in a real world setting with a larger population.

**Methods:** We conducted a retrospective cohort study at “Hospital de Campaña Escuela-Hogar” in Corrientes, Argentina, to assess safety and effectiveness of EPIC in hospitalized adults with severe COVID-19 pneumonia. Primary endpoints were 28-days all-cause mortality and safety. Mortality and improvement in modified WHO clinical scale at 14 and 21 days were secondary endpoints. Potential confounder adjustment was made by logistic regression weighted by the inverse of the probability of receiving the treatment (IPTW) and doubly robust approach.

**Results:** Clinical records of 395 exposed (EPIC) and 446 non-exposed (Controls) patients admitted between November 2020 and April 2021 were analyzed. Median age was 58 years and 56.8% were males. Mortality at 28 days was 15.7% (EPIC) vs. 21.5% (Control). After IPTW adjustment the OR was 0.66 (95 % CI: 0.46 - 0.96), P= 0.03. The effect was more evident in the subgroup who received two EPIC doses (complete treatment, n=379), OR: 0.58 (95% CI 0.39 to 0.85), P=0.005. Overall and serious adverse events were not significantly different between groups.

**Importance:** In this retrospective cohort study, EPIC showed adequate safety and effectiveness in the treatment of hospitalized patients with severe SARS-CoV-2 disease.

## 1. Introduction

From the beginning of the Severe Acute Respiratory Syndrome Coronavirus-2 (SARS-CoV-2) pandemic, different therapeutic approaches have been assessed in randomized controlled trials (RCT) (1). After more than 2 years of pandemic, for hospitalized patients, dexamethasone, remdesivir, tocilizumab, tofacitinib, and baricitinib have shown efficacy in adequately powered clinical trials (2-8). Passive immunotherapy has been widely studied, particularly for patients who have not yet established their specific immune response. To date, convalescent plasma (CP) has shown no effect in the treatment of patients with severe pneumonia. Among ambulatory patients with mild to moderate COVID-19 at high risk for progression to severe disease, neutralizing monoclonal antibodies such as bamlanivimab/etesevimab, casirivimab/imdevimab, and sotrovimab are also included as therapeutic options likewise the administration of nirmatrelvir/ritonavir, 3-day treatment with remdesivir, and molnupiravir (9-12). However, access to these treatments is certainly limited and the appearance and prevalence of the B.1.1.529 variant (Omicron) has threatened the efficacy of most monoclonal antibodies studied to date (13). Effective interventions for the treatment of patients with COVID-19 infection are still urgently needed.

It has been previously shown that the Receptor Binding Domain (RBD) from the viral spike glycoprotein elicits high titers of neutralizing antibodies (NAbs) against SARS-CoV-2 when used as immunogen in horses (14). Equine polyclonal antibodies (EpAbs) represent a practical and efficient source of NAbs. EpAbs are composed of F(ab’)_2_ fragments generated by pepsin digestion. We have formerly described the development of an “Equine Polyclonal Immunotherapy for COVID-19” (EPIC) based on equine anti-RBD F(ab’)_2_ fragments, known as INM005 (15). EPIC showed a very high serum neutralization titer against SARS-CoV-2 and its format, devoid of Fc domains, may prove preferable for its capacity to avoid serum sickness reactions and Fc-triggered side effects (15). The EPIC effect was evaluated in a double-blind, randomized, placebo-controlled trial among hospitalized 241 adults with moderate and severe COVID-19 pneumonia in Argentina (16). Even though the primary endpoint (improvement in at least two categories in WHO ordinal clinical scale at day 28) was not achieved (odds ratio [OR]: 1.61%, 95% confidence interval [95%CI]: 0.71 to 3.63, P=0.34), several secondary endpoints were reached: variation in ordinal clinical status during the follow-up period favored EPIC: A) improvement in at least two categories was significantly higher in the EPIC group at days 14 (OR: 0.52, 95% CI: 0.29 to 0.96, P=0.03), and 21 (OR: 0.54, 95% CI: 0.30 to 0.99, P=0.05); B) a significant difference was noted in time to improvement in at least two ordinal categories or hospital discharge: 14.2 (± 0.7) days in the EPIC group and 16.3 (± 0.7) days in the placebo group (hazard ratio: 1.31, 95% CI: 1.0 to 1.74, P=0.05). Mortality at day 28 was 6.8% in the EPIC group and 11.4% in the placebo group (OR 0.57, 95% IC: 0.24 to 1.37). Pre-specified subgroup analyses showed a more pronounced effect of the intervention among severe patients lacking antibody response at baseline.

With those findings, the Argentinean National Administration of Drugs, Food, and Medical Technology (ANMAT), approved EPIC for the treatment of moderate or severe SARS-CoV-2 hospitalized patients, under special conditions on December 22, 2020 (Provision 9175/20) (17). As required by this approval, the sponsor initiated a prospective nationwide registry of patients with SARS-CoV-2 infection treated with EPIC that has already included more than 10,000 patients. Since data were collected on an ongoing basis, we decided to conduct a retrospective cohort analysis in a dedicated COVID-19 hospital that used the product regularly, including patients admitted to the same hospital before EPIC was available for historical comparison. Therefore, the aim of this study was to evaluate the effectiveness and safety of EPIC administration in hospitalized patients with severe COVID-19 in a real-world setting.

### 2. Objectives

The objectives of the study were to compare all-cause mortality at 28 days and the proportion of serious and total adverse events during hospitalization between the exposed and unexposed groups.

### 3. Study Design and location

A retrospective cohort study was performed including adult patients hospitalized for severe pneumonia with confirmed diagnosis of SARS-CoV-2. The date of hospital admission was considered as the date of admission into the cohort for both groups. All patients were evaluated in a structured manner at days 14, 21 and 28 from inclusion.

All cohort participants were enrolled at the “Hospital de Campaña Escuela-Hogar” (HCEH), a single monovalent site established as a reference center for the diagnosis and treatment of patients with COVID-19 for the whole province in Corrientes city, located in the northeastern region of Argentina. HCEH is a paper-free facility with 723 beds, 300 of which are equipped for critical care.

The study cohort subjects were divided in two groups: those “exposed to EPIC or EPIC group” that received treatment with purified equine anti-RBD F(ab’)_2_ fragments (EPIC, CoviFab®) and those “Non-exposed or Control group” that never received that intervention.

Following ANMAT’s approval, issued on January 27th, 2021, EPIC was adopted as standard of care for patients with COVID-19 within the HCEH. Therefore, we selected that date to divide the cohort groups in exposed cases onward from that date to April 17th, 2021, and non-exposed cohort group backward from January 21st, 2021 to November 25th, 2020 in order to increase comparability, and also to avoid the possibility of a learning curve period for the treatment of patients with COVID-19 close to the beginning of operations at HCEH.

The study protocol was approved by the Institutional Review Boards from both the HCEH and the “Hospital Italiano” of Buenos Aires city, Argentina. The protocol was registered in ClinicalTrials.gov with the number NCT04954235. All data extracted from the electronic clinical records was anonymized at completion of the structured study forms and remained in that way for the whole analysis process.

## 4. Methods

### Population

Adult patients with confirmed diagnosis of SARS-CoV-2 severe pneumonia hospitalized at HCEH between November 25th, 2020 until April 17th, 2021, meeting the study selection criteria.

### Exposure

Patients were labeled as exposed (EPIC) if they received at least one of 2 doses of 4 mg/Kg of EPIC in intravenous infusion of 100 ml of saline over a period of fifty minutes (with an interval of 48 hours between doses). The control group was recruited among hospitalized patients that did not receive that intervention because they were hospitalized before EPIC was available.

### Selection criteria

Patients were included in the study if had between 18 and 79 years old, COVID-19 diagnosis confirmed by SARS-CoV-2 antigen test or reverse-transcriptase-polymerase-chain reaction (qRT-PCR -GeneDX Co, Ltd or similar) or positive anti SARS-CoV-2 IgM antibodies, had severe disease defined as: respiratory rate of more than 30/min, or oxygen saturation <94% on room air at sea level, or a ratio of arterial partial pressure of oxygen to fraction of inspired oxygen (PaO2/FiO2) <300 mm Hg, or lung compromise of more than 50%.

Exclusion criteria were: SARS-CoV-2 disease other than severe (asymptomatic, mild, moderate or critical), pregnant women or during lactation period, patients already admitted to intensive care unit (ICU), confirmation of microbiological cause of pneumonia other than SARS-CoV-2, patients with therapeutic limitation or patients with history of anaphylaxis or severe allergic reaction to equine sera or to contact or exposure to horse proteins. A detailed description of the disease severity categories is provided in the Supplementary Appendix.

### Data collection

Data capture was performed through ACTide-eCRF, an electronic case report form based on a multi-platform interface that was previously reviewed by ANMAT and used for the registry of all patients receiving EPIC in accordance with Provision 4622/12 regarding “Authorization under Special Conditions”. This platform was in full agreement with current regulations regarding validation, compliance and certification of computing systems (e.g., CFR 21 Part 11, EU GMP Annex 11, GAMP5, GDPR and HIPAA). A specific allowance system was implemented for the study based upon personnel roles.

Patients from the exposed group provided written informed consent for the administration of EPIC. A team of physicians trained on data collection retrieved the required information for each selected patient at baseline, and at days 14, 21, and 28 using specifically designed structured forms.

Patients from the non-exposed group were identified through a systematic review of the electronic clinical records from HCEH, and their data was retrieved by the same data collection team using the same structured forms.

### Variables

Structured forms used for both cohort groups included demographic variables (age in years, gender at birth and body mass index-BMI), comorbidities (hypertension, diabetes, obesity, cancer, chronic obstructive pulmonary disease, chronic hepatic insufficiency, chronic renal insufficiency and cardiovascular disease), use of convalescent plasma since 30 days prior to hospital admission, Charlson Score as a measure of comorbidity prognosis, clinical variables at baseline (diagnostic method, time from symptoms onset, respiratory rate, oxygen saturation, PaO2/FiO2, requirement of supplementary oxygen and National Early Warning Score-NEWS), and variables related to clinical outcomes including WHO-modified 6-points ordinal clinical scale (18-20). Further details of variable operationalization are provided in the Supplementary Appendix.

### Outcomes

Primary outcome was all-cause mortality between cohort groups at day 28 since cohort admission. Secondary outcomes included mortality between groups at day 14 and 21 since cohort admission, WHO-modified 6-points ordinal clinical scale (1 non-hospitalized with full restitution of physical functions, 2 non hospitalized without complete restitution of physical functions, 3 hospitalized without oxygen requirement, 4 hospitalized requiring supplemental oxygen, 5 invasive ventilatory support and 6 death) measured at days 14, 21 and 28, the proportion of patients discharged from hospital at same intervals, time (in days) between cohort admission and hospital discharge, proportion of patients transferred to ICU, time (in days) since cohort admission to discharge from ICU, proportion of patients requiring mechanical ventilation, and time (in days) since cohort admission to start of mechanical ventilation (21).

Safety outcomes included the percentage of patients suffering from any type and/or serious adverse events. For all adverse events the following data was captured: start and end date, severity, causality assessment, and treatment. The Adverse Event table was organized according to the MedDRA SOC (System Organ Class).

### Sample size

Based upon a prior experience with the use of EPIC in the context of a clinical trial, an estimated mortality of 24% for the “non-exposed” patients was assumed. As such, a reduction in mortality of at least 8% (e.g., 16% for exposed and 24% for non-exposed patients) was considered clinically meaningful. With these assumptions, a minimal inclusion of 392 exposed and 392 non-exposed patients (784 patients in total) was required for a two-tailed test with an a error of 5% and 80% of statistical power. We aimed to include a sample size of 800 patients (1:1 ratio with 400 in each cohort group).

### Statistical methods and bias management

We used absolute and relative frequencies (percentages) to describe categorical variables, mean and standard deviation (SD), or median and interquartile range (IQR: 25th centile and 75th centile) to describe quantitative variables. We used Chi-square test or Fisher’s exact test for categorical variables and T test or Wilcoxon rank sum test to compare the exposed and non-exposed characteristics.

Effectiveness estimates for treatment might be confounded by indication as a result of treatment decisions based on observed characteristics. Given the observational nature of the study, we use an inverse probability of treatment weighting (IPTW) approach to estimate the causal association -also referred as average treatment effect-of EPIC considering potential confounders (22). This method, which is an extension of the propensity score, is particularly helpful in the estimation of causal associations when a randomized clinical trial is unfeasible. The IPTW approach generates weighted cohorts removing the differences in relevant observed confounders (23).

We estimated the propensity score (PS) of EPIC exposure using a logistic regression model with EPIC exposure as dependent variable and the following potential predictors of treatment: gender at birth, age, clinical parameters at cohort admission (respiratory rate, heart rate, body temperature, oxygen saturation), requirement of supplementary oxygen or non-invasive ventilation, Charlson’s Score, National Early Warning Score (NEWS), time from symptoms onset, prior use of angiotensin converting enzyme inhibitors, non-steroidal antiinflammatory agents, corticosteroids, heparin, immunosuppressants, ivermectin or statins; presence and number of comorbidities: obesity, cardiovascular disease, stroke, hemiplegia, hypertension, chronic lung disease, chronic renal disease, dementia, peptic ulcer, diabetes with or without target organ damage, solid organ tumor or leukemia. With this propensity score we calculated the stabilized IPTW. The weights were truncated at percentile 1% and 99% to avoid extreme figures.

We used Cox’s proportional hazards regression model to estimate the hazard ratio (HR) for calculation of the primary endpoint. We also informed the OR between cohort groups calculated with a logistic regression model. The effect of EPIC on the distribution of the modified WHO-clinical ordinal scale at 14, 21 and 28 days was accessed with an ordinal logistic regression model and presented as proportional Odds Ratio using Brant test (parallel regression assumption test) (pOR) (24-28). We used conventional logistic regression models to estimate the Odds Ratio (OR) of the exposure to EPIC on the comparison of the proportion of hospital discharge at 14, 21 and 28 days, and the proportion of adverse effects. We use Fine and Gray’s regression model to estimate the subhazard ratio (sHR) for the association between EPIC and the requirement of mechanical ventilation using death as a competing event (29).

For the primary analysis we estimated the causal association between EPIC exposure and primary or secondary outcomes using regression models weighted by the inverse of the probability of receiving treatment, considering EPIC exposure as the independent variable (IPTW). In addition, we presented the data with a conventional adjustment for potential confounders either with the raw data and with the IPTW-treated data, in a doubly robust approach. Variables included in this adjustment included: gender at birth, age, respiratory rate at cohort entry, oxygen saturation, requirement of supplementary oxygen or non-invasive ventilation, Charlson’s Score, NEWS, time from symptoms onset, obesity, cancer and chronic lung disease. We presented all data with 95% confidence interval (95% CI). To assess the balance in potential confounders between exposure groups, we used standardized differences before and after IPTW adjusting. We considered balanced covariates all those who had standardized differences lower than 10% after IPTW adjusting (30). We presented the balance diagnosis of IPTW in a Love plot according to Austin and Stuart (31).

We performed predefined subgroup analysis of primary outcomes considering gender at birth, age category groups (less than 65, or between 65 and 79 years old), time from symptoms initiation (less than 3 days, between 3 and 5, between 5 and 10 or more than 10), obesity, presence and number of main comorbidities (immunosuppression, diabetes, hypertension, cardiovascular disease) and number of doses of EPIC (one versus two). We also performed a sensitivity analysis considering the patients from the “Control” group that did or did not receive convalescent plasma.

The detailed statistical analysis plan is provided in the Supplementary Appendix. All tests were two-sided, and a P value <0.05 was considered statistically significant. All statistical analysis was performed using STATA statistical software version 15.1 MP - Parallel Edition (Copyright 1985-2017 StataCorp LLC - StataCorp. 4905 Lakeway Drive, College Station, Texas 77845 USA).

## 5. Results

### Patients

Following protocol’s approval, data collection from both cohort groups started on May 27th, 2021. During the pre-specified -backward and onward-collection times, from 1352 clinical records reviewed, 395 patients met the inclusion criteria for the EPIC group and 446 patients were included in the Control group, for a total cohort of 841 patients. Fig. 1 depicts the complete patient disposition.

**FIG 1.**
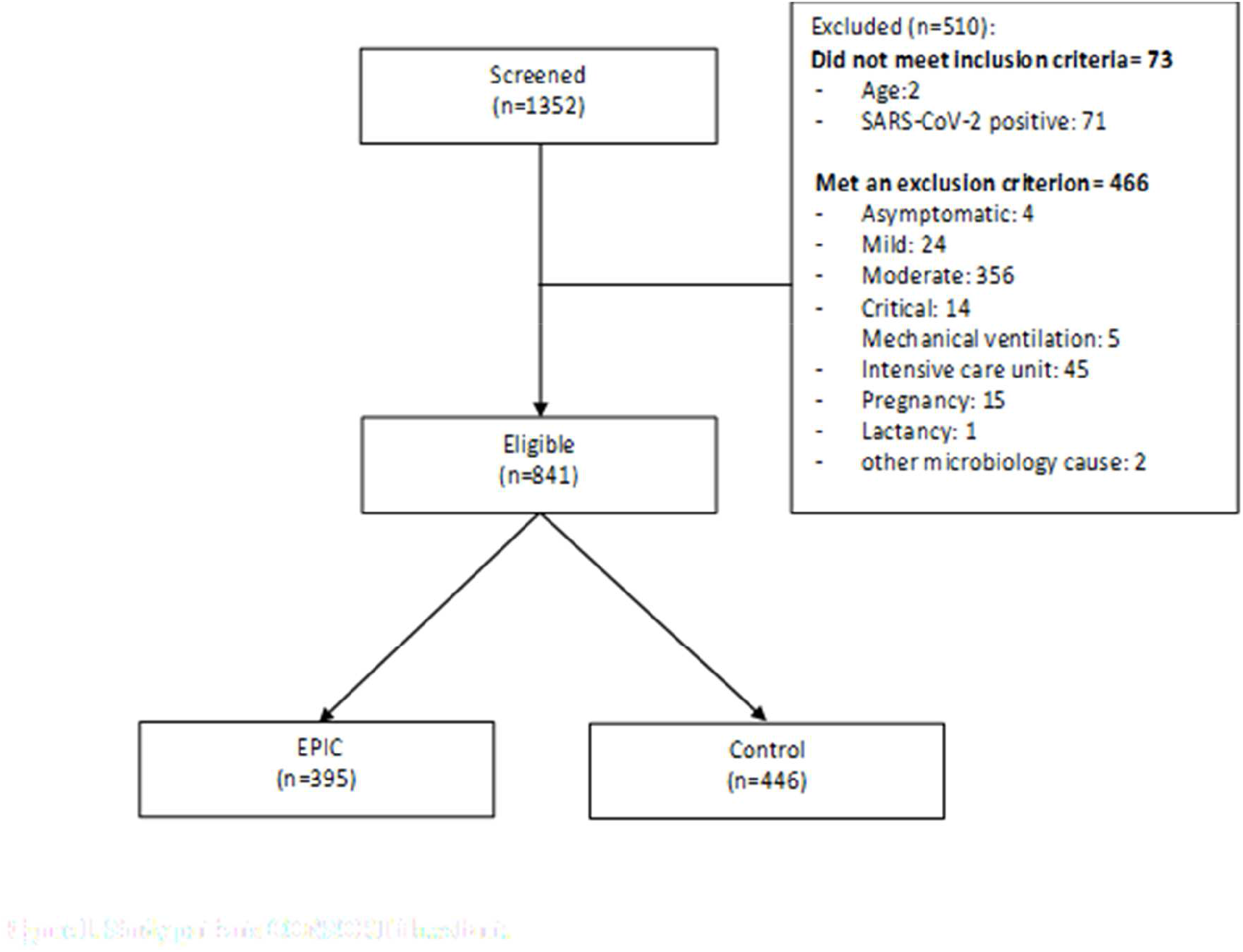
Cohort patient disposition

Median age was 58 years old (interquartile range 48 to 69), and 57.9% were male. Body mass index was 31.1 (interquartile range 28.1 to 36) in the EPIC group and 28.6 (interquartile range 26 to 33.2) in the Control group, with 117 and 235 patients in the EPIC group, and 194 and 176 patients in the Control group reaching the standard definition for overweight and obesity, respectively. Standard and modified Charlson’s scores at cohort entry were similar between groups.

At cohort admission the median time from symptoms initiation was 7 days (interquartile range 5 to 8) for the EPIC group and 6 days (interquartile range 3 to 9) for the Control group.

While most patients had at least one identified comorbidity, significantly more patients had no comorbidities in the Control group than in the EPIC Group: 67 (15%) vs. 36 (9.1%), respectively, p=0.009. Obesity -235 patients (59.5%) and 176 (39.5%), P=<0.001-along with hypertension - 228 (57.7%) and 242 (54.3%)-, and diabetes -111 (28.1%) and 110 (24.7%) in EPIC and Control groups, respectively, were the most prevalent coexisting conditions.

No statistically significant differences were found in body temperature, and heart and respiratory rates between cohort groups, whereas oxygen saturation at cohort entry was 92% (interquartile range 89 to 93%) for EPIC group and 90.5% (interquartile range 88 to 93) for the Control group, (P<0.0001). Accordingly, the modified NEWS score at cohort admission was significantly different between groups (P=0.006).

A total of 46.3% of the cohort patients (179 in EPIC group and 211 in the Control group) were receiving antihypertensive drug treatment (angiotensin converting enzyme inhibitors and angiotensin receptor blockers) at admission, while a minority of patients received other drugs as immunosuppressants, statins, anticoagulants, ivermectin, and corticosteroids, none of them being statistically different between groups. A more detailed description of the baseline patient conditions and comparison between cohort groups is shown in Table 1.

**TABLE 1.** Demographic and clinical characteristics of the patients at baseline

|  | EPIC<br>(N=395) | Control<br>(N=446) | P value |
| --- | --- | --- | --- |
| Characteristics |  |  |  |
| Age, years# | 57 (48-67) | 59 (48-69) | 0.1881† |
| Male sex | 60.3 (238) | 55.8 (249) | 0.195 |
| BMI** |  |  |  |
| Median (Interquartile range)# | 31.1 (28 - 36) | 28.6 (26-33.2) | <0.0001† |
| Normal | 10.9 (43) | 12.6 (56) | <0.001 |
| Overweight | 29.6 (117) | 43.5 (194) |  |
| Obesity | 59.5 (235) | 39.5 (176) |  |
| Coexisting conditions |  |  |  |
| Charlson score | 0 (0 - 2) | 0 (0 - 2) | 0.9982† |
| Number of comorbidities# | 2 (1 - 3) | 1 (1 - 3) | 0.0003† |
| None | 9.1 (36) | 15.0 (67) | 0.009 |
| Hypertension | 57.7 (228) | 54,3 (242) | 0.313 |
| Diabetes | 28.1 (111) | 24.7 (110) | 0.258 |
| Obesity | 59.5 (235) | 40.6 (181) | <0.001 |
| Lung disease | 7.1 (28) | 10.5 (47) | 0.080 |
| Baseline characteristics at time of hospital admission |  |  |  |
| Days from symptoms onset to hospital admission# | 7 (5 - 8) | 6 (3 - 9) | 0.0014† |
| Hospitalized, requiring supplemental oxygen | 95.2 (376) | 93.5 (418) | 0.355 |
| Hospitalized, receiving noninvasive mechanical ventilation or high-flow oxygen devices | 0 (0) | 0.7 (3) | 0.252‡ |
| Oxygen saturation on room at cohort entry | 92 (89 - 93) | 90.5 (88 - 93) | 0.0014† |
| Respiratory frequency | 20 (20 - 21) | 20 (20 - 22) | 0.3096† |
| Heart rate | 89 (80 - 100) | 90 (78 - 102) | 0.3703† |
| Temperature | 36.3 (36 - 37) | 36.2 (36 - 36.9) | 0.4612† |
| NEWS |  |  |  |
| Low | 36.0 (142) | 28.3 (126) | 0.006 |
| Medium-low | 19.8 (78) | 19.3 (86) |  |
| Medium | 34.7 (137) | 35.4 (158) |  |
| High | 9.6 (38) | 17.0 (76) |  |
| Concomitant medications prior to cohort admission |  |  |  |
| ACE inhibitors / AIIRA | 45.3 (179) | 47.3 (211) | 0.563 |
| NSAIDs | 17.0 (67) | 15.3 (68) | 0.499 |
| Immunosuppressors | 1.5 (6) | 2.2 (10) | 0.444 |
| Statins | 8.9 (35) | 8.1 (36) | 0.681 |
| Ivermectin | 6.3 (25) | 3.4 (15) | 0.044 |
| Anticoagulants | 3.8 (15) | 4.0 (18) | 0.859 |
| Corticosteroids | 8.6 (34) | 11.0 (49) | 0.248 |

\*\* Data was missing from 20 patients in Control Group
All data are % (n), except for: # Data are in median (interquartile range <sub>25:75</sub>)
All p values are for Chi2 except † for Wilcoxon rank-sum (Mann-Whitney) test, and ‡ for Fisher's exact test

During the cohort follow up time 395 patients (100%) in the EPIC group and 406 patients (91%) in the Control group received corticosteroids, 392 (99%) and 410 (91.9%) received heparin, 274 (69.4%) and 321 (72%) received antibiotics, respectively, while 317 (71.19%) in the Control group received convalescent plasma (exclusion criterion for EPIC group). A complete description of concomitant treatment interventions during cohort follow up is shown in Table 2.

**TABLE 2.** COVID-19 treatment intervention during hospitalization

|  | **EPIC**<br>(N=395) | **Control**<br>(N=446) | **P value** |
| --- | --- | --- | --- |
| **Concomitant medication during hospitalization** |  |  |  |
| **None** | 0 (0) | 0.7 (3) | 0.252<sup>‡</sup> |
| **Corticoids** | 100 (395) | 91 (406) | <0.001 |
| **Heparin** | 99.2 (392) | 91.9 (410) | <0.001 |
| **Convalescent plasma** | 0 (0) | 71.1 (317) | <0.001 |
| **Monoclonal antibodies** | 0.3 (1) | 0 (0) | 0.470<sup>‡</sup> |
| **Antibiotics** | 69.4 (274) | 72 (321) | 0.407 |
| **Ivermectin** | 0 (0) | 0.2 (1) | 1.000<sup>‡</sup> |
| **Tocilizumab** | 0 (0) | 0.2 (1) | 1.000<sup>‡</sup> |
| **Colchicine** | 0 (0) | 0.2 (1) | 1.000<sup>‡</sup> |
| **Remdesivir** | 0.3 (1) | 0 (0) | 0.470<sup>‡</sup> |

All data are % (n)
All P values are for Chi2 except † for Wilcoxon rank-sum (Mann-Whitney) test, and ‡ for Fisher's exact test

### Intervention

No patients in the Control group received polyclonal specific hyperimmune serum against SARS-CoV-2. All three hundred and ninety-five patients in the EPIC group received one dose of hyperimmune serum and 379 patients (96%) completed the two-dose pre-specified treatment. Reasons for failure in completing the intervention included admission to UCI and staff refusal to continue treatment (10 cases), adverse events (2 cases), patient voluntary discharge from Hospital (1 case), patient refusal (1 case) and other reasons (2 cases).

### Primary outcome

Overall mortality at 28 days was significantly lower for patients in the EPIC group than for patients in the Control group. The OR was 0.66 (95% CI 0.46 to 0.96 p=0.03) and the HR for the IPTW-adjusted data was 0.70 (95% CI 0.50-0.98 p=0.037). The doubly robust approaches provided similar results (Table 3, Fig. 2).

**TABLE 3.**
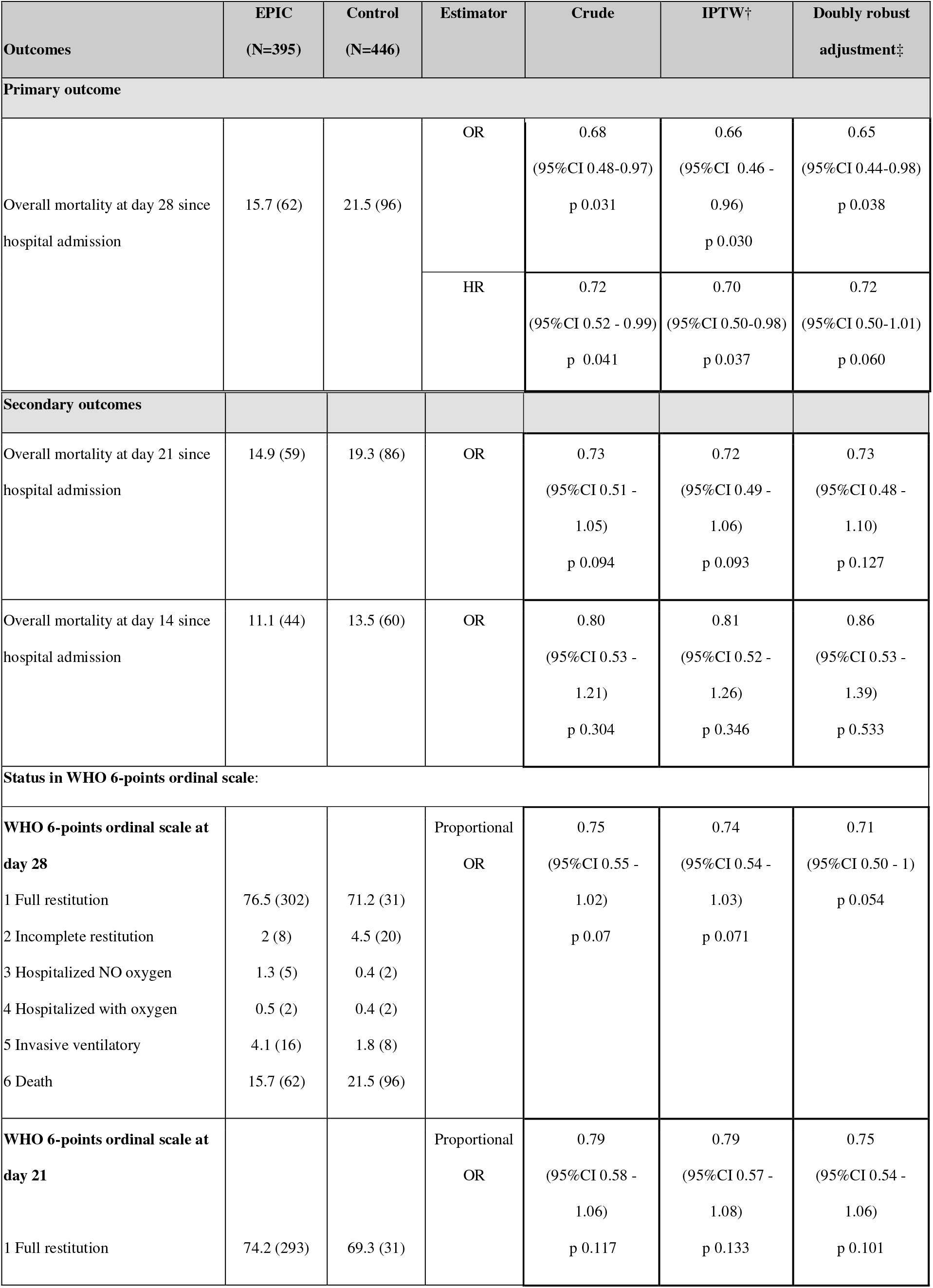
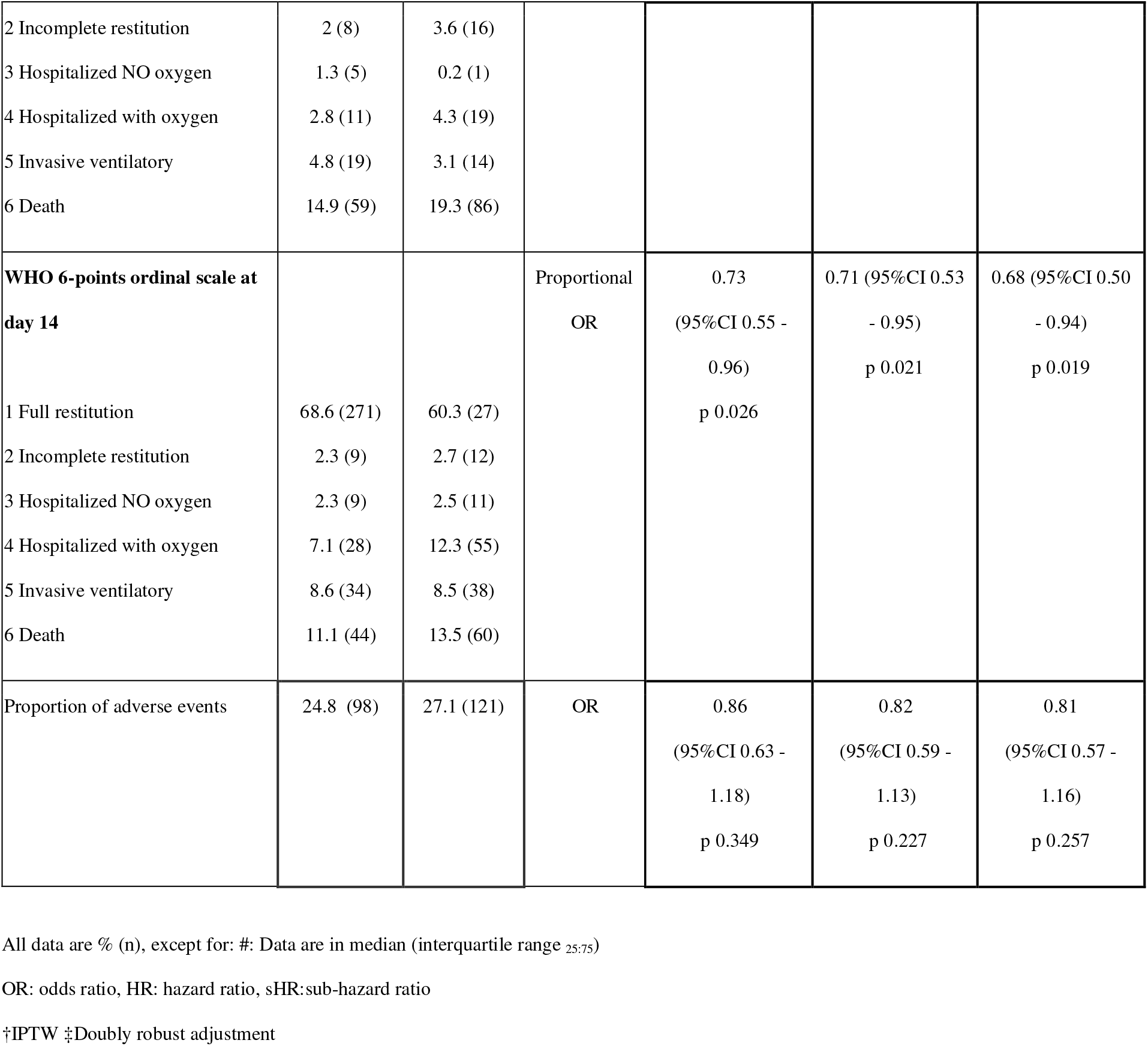
Clinical Outcomes in EPIC and control groups

**FIG 2.**
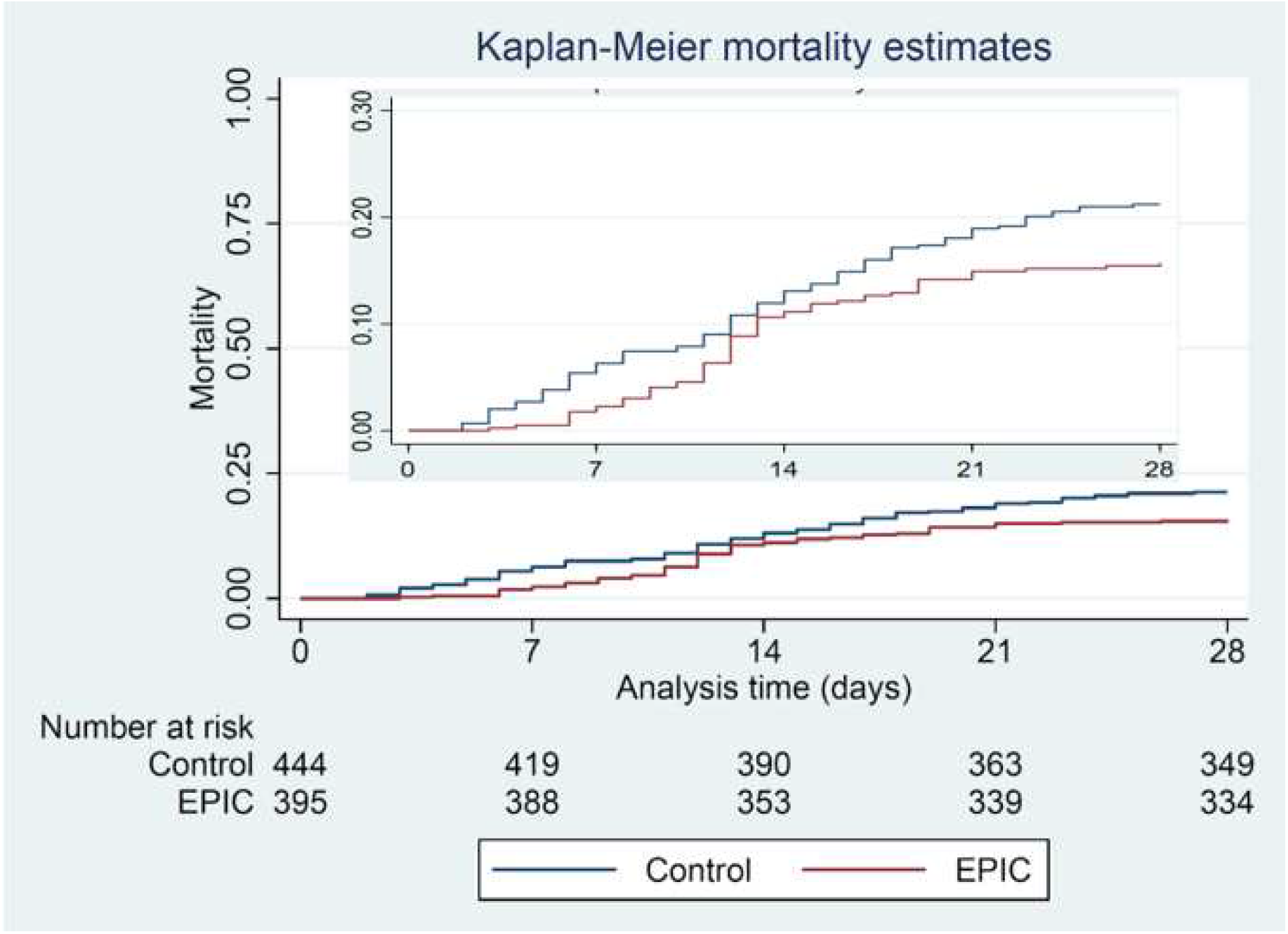
Overall mortality at day 28 since cohort admission

### Secondary outcomes

Overall mortality at 21 and 14 days were not statistically different between cohort groups (OR for the IPTW-adjusted data and for the doubly robust approach were 0.72 [95% CI 0.49 to 1.06] and 0.73 [95% CI 0.48 to 1.10], and 0.81 [95% CI 0.52 to 1.26] and 0.86 [95% CI 0.53 to 1.39], respectively.

WHO-modified 6 points ordinal clinical scale results were significantly better for the EPIC group than for the Control group at 14 days (proportional OR 0.71 [95% CI 0.53 to 0.95] for IPTW-adjusted and proportional OR 0.68 [95% CI 0.50 to 0.94] for the doubly robust approach), while no significant differences were noted at days 21 and 28 between groups (Table 3, Fig. S2).

Hospital discharge rate was also significantly higher for the EPIC group than for the Control group at 14 days (OR 1.46 [95% CI 1.07 to 1.98] for IPTW-adjusted and OR 1.49 [95% CI 1.07 to 2.09] for the doubly robust approach), while no differences were found between cohort groups at days 21 and 28.

No significant differences were found between groups in median time to hospital discharge, proportion of patients requiring admission to ICU, median time until discharge from ICU, proportion of patients requiring mechanical ventilation or median time until initiation of mechanical ventilation. Complete results for secondary outcomes are shown in Table S2 and Fig. S3, S4 and S5.

### Safety outcomes

Safety outcomes included the percentage of patients with any type and/or serious adverse events, and the comparison of total and serious adverse events (AE) during hospital admission between EPIC and Control groups.

EPIC was overall well tolerated, 98 patients (24.8%) in the EPIC group and 121 (27.1%) in the Control group had at least one adverse event, being respiratory, thoracic and mediastinal disorders (20.3% in EPIC group and 26% in Control group), and administration site complications (11.6% in EPIC and 9.6% in Control group) the most frequently observed, with no statistical difference between cohort groups. Two patients out of 395 in the EPIC group did not receive the second dose of the study product due to adverse events, i.e., a cutaneous rash and a hypotension event, both of which resolved satisfactorily.

Serious adverse events (SAE) were frequently observed among cohort patients, probably due to the population characteristics, although no significant difference between groups was observed. SAE occurred in 78 patients (19.7%) in the EPIC group and in 106 (23.8%) in the Control group. Six AEs were considered probable or possibly related to EPIC.

No anaphylaxis event was reported, and no death episodes were considered by the investigators to be related to the investigational product. Table 4 and Table S4 show a more detailed description of the AE between cohort groups.

**TABLE 4.** Review of safety outcomes

|  | **EPIC**<br>**(N=395)** | **Control**<br>**(N=446)** |
| --- | --- | --- |
| **Any patient with an adverse event** | 24.8 (98) | 27.1 (121) |
| **Total of adverse events** | 145 | 168 |
| **Type** |  |  |
| Mild AE | 1.8 (7) | 0.2 (1) |
| Moderate AE | 2.3 (9) | 1.6 (7) |
| Severe AE | 12.9 (51) | 12.1 (54) |
| Serious adverse events | 19.7 (78) | 23.8 (106) |
| **Relatedness** |  |  |
| Not related AE | 35.2 (139) | NA |
| Possible related AE | 0.8 (3) | NA |
| Probable related AE | 0.8 (3) | NA |
| **Association** |  |  |
| Associated to concomitant disease | 22 (87) | 18.4 (82) |
| Associated to concomitant medication | 0.5 (2) | 0 |
| Associated to COVID-19 | 33.4 (132) | 35.6 (159) |

NA: Not applicable

## 6. Discussion

In this retrospective study, the administration of at least one infusion of EPIC for hospitalized patients with severe COVID-19 significantly reduced the 28-day mortality by 34% (15.7% vs 21.5%) compared with a group of patients treated at the same hospital before EPIC was available. This effect was more evident in patients receiving the two infusions of the investigational product (28-day mortality of 14% vs 21.5%), resulting in 13 patients needed to treat in order to prevent one additional death. During the trial, EPIC-related adverse events were minimal and non-serious.

Currently, no virus directed therapy has shown to reduce mortality among hospitalized patients with severe SARS-CoV-2 infection; only agents decreasing the inflammatory response such as dexamethasone, baricitinib (or tofacitinib), and tocilizumab (or sarilumab) were able to reduce the mortality rate in this setting (3, 7, 32, 33).

Administration of different types of immunotherapy has been explored as a therapeutic approach for patients with severe COVID-19 disease with variable results. Recent data from three randomized clinical trials studies comparing anti-spike mAbs (bamlanivimab, VIR-7831, and the combination BRII-196/198) with placebo were terminated due futility after randomization of 314, 344 and 343 patients, respectively (34, 35).

However and as expected, most of the clinical benefits obtained from passive immunotherapies might be observed in patients lacking specific anti-SAR-CoV-2 antibodies due to a delayed response or to immunocompromise. In this regard, among patients randomized to placebo in the Recovery trial, the 28-day mortality was significantly higher in the baseline seronegative group (30%) than in the seropositive one (15%). Even more, in those lacking specific antibodies at baseline, the mortality was 24% and 30% in the mAbs combination and placebo arm, respectively (rate ratio 0.80) (36).

The positive impact of polyclonal equine anti-RBD F(ab’)_2_ fragments described here might follow the findings reported in the Recovery clinical trial. Given the observational nature of this study, baseline anti-SARS-CoV-2 immune status could not be determined. Lacking this information forbidden us to identify a subgroup of seronegative patients who might have benefited the most from this type of therapy. Remarkably, a significant effect on the primary outcome was observed for the entire population while in the Recovery trial that enrolled 9785 patients, a decrease in the 28-day mortality was seen only in those with no baseline antibodies (36).

It should be noted that, in a double-blind randomized clinical trial, the treatment of EPIC was associated with a 45% non-statistically significant decrease in mortality among the subgroup of severe patients hospitalized with COVID-19 disease (16) (13.6% in the EPIC group vs 24.5% in the placebo group, OR: 0.52). It is likely that this outcome did not reach statically significance because of the low number of severe patients enrolled. Moreover, EPIC showed a statistically significant difference in the WHO-modified 6 points ordinal clinical scale at 14 days. This was correlated with also a statistically significant difference at hospital discharge rate at 14 days. These effects were not observed neither at day 21 nor at day 28 (16).

In regard to safety results, these findings were as expected in patients with COVID-19 severe disease as adverse events observed were mainly related to disease progression and complications such as multiorgan failure and in similar proportions in both groups. EPIC showed a good safety profile similar to the one observed in the RCT. All related events were treated and resolved satisfactorily within the same date. In this real-world experience only in 2 of the 395 patients treated with EPIC the second dose was not infused due to adverse events during the first dose. These findings are aligned with the adverse event reporting rate observed through the Argentinean EPIC registry of 10.728 patients where hypersensitivity events were reported in a frequency of 1-2% through the period December 2020 and August 2021 (37).

The main limitation of the study is the non-randomized design; however, data collection included structured forms applied for subjects enrolled in both cohort groups and at the same Hospital. Even more, all hospitalized subjects with COVID-19 were evaluated and enrolled in the study in a consecutive manner. Another existing limitation is that patients were included in one clinical site, missing the generalizability observed in multicenter trials. Of note, the results shown here should not be extrapolated to the vaccinated population nor to those with prior COVID-19 disease since only one participant had received a COVID-19 vaccine and a history of SARS-CoV-2 infection was an exclusion criterion.

Although this was not fully studied, it can be speculated that most of the patients enrolled in this study were probably infected with the Gamma or P.1 variant due to the epidemiological situation of Argentina at the time of study. Nevertheless, being of polyclonal nature, EPIC has a broader recognition of epitopes on RBD Spike protein than mAbs, making this immunotherapy more robust against viral scape mutations. Interestingly, EPIC showed high neutralizing activity against Gamma and Delta variants of concern and was also capable of neutralizing Omicron, although with less efficiency (Gallego S. et al., unpublished data). Therefore, the clinical benefit observed in this study could be extrapolated to patients infected with any of the above-mentioned variants of concern.

## 7. Conclusion

After proper adjustment by confounders, this retrospective study in a real-world setting suggests a favorable effect of EPIC on the 28-day mortality rate in patients with severe SARS-CoV-2 infection admitted to a monovalent hospital site in Argentina, with an adequate safety profile.

## Data Availability

All data produced in the present study are available upon reasonable request to the authors

https://ClinicalTrials.gov

## Funding

Funded by Inmunova SA. and Laboratorio Elea Phoenix S.A.

## Notes

### Competing Interest Statement

DHFS, FA, AAG, NAO, SNMP, LSN, FC, OIMI declare reimbursement for conduction of the observational study as investigators. EN, GL, WHB, DHG and LP report personal fees from Inmunova. SG and BK declare no competing interests.

### Clinical Protocols

https://ClinicalTrials.gov

### Funding Statement

Inmunova SA and Laboratorios Elea-Phoenix SA

### Author Declarations

The study protocol was approved by the Institutional Review Boards from both the Hospital de Campanha Escuela-Hogar and the Hospital Italiano of Buenos Aires city, Argentina.

